# Impact of booster COVID-19 vaccine for Moroccan adults: A discrete age-structured model approach

**DOI:** 10.1101/2021.03.14.21253555

**Authors:** Aayah Hammoumi, Hanane Hmarrass, Redouane Qesmi

## Abstract

Public health control strategies, such as lockdown, seem to be effective in reducing the spread of the pandemic, but are ineffective as a whole since lockdown is responsible of global economic crisis and badly lived by the majority of children and adults who have developed mental health disorders and familial problems as well. Thus, the development of a vaccine against COVID-19 is needed to control this disease. We have developed a discrete age-structured model and followed the Moroccan vaccination program to assess the impact of booster vaccination targeting Moroccan adults against COVID-19. Using the derived model, we estimated some relevant model parameters related to COVID-19 using collected cumulative mortality and reported Moroccan data. A control reproduction number *R*_*c*_, which determines the necessary level of vaccine uptake that lead to COVID-19 eradication is determined. Furthermore, a herd immunity threshold above which the population can be protected from COVID-19 infection is derived. Analyzing the model, sufficient and necessary conditions for the eradication of the disease are obtained as well. Next, we perform numerical simulations to study the impact of several uptake levels of the potential vaccine on the persistence and the extinction of COVID-19 pandemic. Our results show that the COVID-19 is expected to last past 2021 in the absence of a vaccination program. Moreover, a vaccination of the adult population at rate 0.6% per day needs at least 67% of vaccine efficacy and 90% of immunogenicity rate to eradicate the disease. Using Sinopharm vaccine, the herd immunity can be achieved when about half of Moroccan adult population is immunized against the COVID-19. However, using Oxford-Astrazeneca vaccine, less than 60% of adult population must be immunized against the disease to achieve the herd immunity. Finally, if vaccine efficacy is about 80% and the immunogenicity is about 50% then vaccinating adults at rate of 0.6% per day could protect roughly 22% of children from COVID-19 infection.

## 1. Introduction

The newly emerging coronavirus diseases become constant threats to global public health [50]. There are seven coronaviruses (CoVs) known to infect humans so far [63]. Among them, three have caused more severe respiratory illness and fatalities including SARS-CoV, MERS-CoV and SARS-CoV-2 [50]. Over the two past decades, they were responsible for three major coronaviruses outbreaks. Indeed, in November 2002, the first case of severe acute respiratory syndrome (SARS) emerged in Guangdong, China. The disease has reached pandemic by July 2003 and mysteriously disappeared in the same year with a total of 8,096 reported cases, including 774 deaths in 27 countries [43]. Ten years later, the Middle East respiratory syndrome (MERS) emerged in Saudi Arabia and caused an epidemic in the Middle East. Between April 2012 and December 2019, there have been 2499 confirmed cases of MERS and 858 deaths in 27 countries [23, 35]. The most severe acute respiratory syndrome coronavirus 2 (SARS-CoV-2) is the last coronaviruses which probably emerged in China in December 2019 and continues to progress to date [18, 56]. Although less lethal than SARS and MERS, SARS-CoV-2 is more transmissible and, the so-called COVID-19 disease, reached a pandemic status in mid-March 2020 [50, 18, 10]. Affecting 218 countries and territories around the world, it is responsible actually of more than 61 million cases of infection and 1.4 million deaths globally. Furthermore, despite the public health measures taken to control the disease spread, the world is facing an unprecedented economic crisis and the vaccination becomes the only hope to eradicate the COVID-19 and to allow people to return to normal life [39]. An effective COVID-19 vaccine must provide a strong and long term protective antibody response [11]. Unfortunately, until the end of 2020, there were no vaccine approved against any Human coronaviruses. In the past, several vaccine candidates for SARS and MERS have been under development but the research program has been shelved because of three reasons [2, 13]. First of all, the natural extinction of SARS-CoV occurred before the process of vaccine development was completed. Secondly, there was no enough financial support for vaccine development, and, finally, the market size deemed to be too small by the large pharmaceutical companies [56, 6]. However, due to the great genetic similarity between SARS-CoV and SARS-CoV-2, the acquired knowledge is now very useful for COVID-19 vaccine development [55]. Different technology platforms are performed including both traditional (i.e. inactivated virus, live attenuated virus and protein/adjuvant) and novel approaches (i.e. viral vectors and nucleic acids) [29, 52, 47]. Furthermore, supported by WHO and partners, several pharmaceutical companies and academic institutions worldwide joined their effort to produce a safe and effective vaccine as soon as possible in early 2021 [47, 59, 64]. This constitutes a real challenge as vaccine development process takes more than a decade [15]. In the case of SARS-CoV-2, the internationally race for COVID-19 vaccines started shortly after first sequencing of its complete genome by China in January, 2020 [22]. Currently, at least 151 vaccine candidates are in preclinical evaluation, and about 48 are in clinical evaluation for which a dozen are available or nearing the final stage of testing [60, 37, 47]. Once approved, the WHO and partners will guarantee access to these vaccines worldwide though the COVAX program in order to cover 20 of the world’s population [60, 61]. Health workers and vulnerable people will be prioritized. Additional doses of vaccine may be provided if the epidemiological situation of country so requires [61]. Morocco is one of the countries hitted by COVID-19 with a total of 428193 confirmed cases and 7170 deaths as of December 26*th*; 2020; [48]. The Moroccan government decided then to be among the countries to conduct a mass emergency vaccination campaign against the COVID-19. Since the end of December 2020, the Moroccan Kingdom intends to launch an emergency vaccination program for the adult population upon immediate receipt of the first pre-ordered doses of vaccines from the Sinopharm or Astrazeneca companies. It will cover 80% of the population aged over 18 years old. The program will be conducted over a period of 12 weeks in which people will receive two doses of vaccine separated by 21 days in average.

Mathematical modeling has proven to be a powerful tool to predict the dynamics of the epidemic and to help decision makers to evaluate the effectiveness of public health control interventions (See [31, 38, 53, 19] and references therein). Furthermore, mathematical models can be used to assess whether a vaccination strategy will be effective in eradicating or controlling a disease. Using a mathematical model of COVID-19 vaccine portfolio in [34], McDonnell et al. estimated timelines and probabilities of success of COVID-19 vaccines. Their results suggests that, by the end of April 2021, there is a 50% of chance that a safe and efficient vaccine can be approved by the end of 2021. Furthermore, it could take more than a year to produce sufficient vaccines to cover 50 million medical staff in the entire world. In [26], Peter Jentsch et al. developed an age-structured SEAIR model of COVID-19 with vaccination in Ontario to check whether it is better to vaccinate the most vulnerable individuals or those who spread the virus further to others. They found that using vaccines to break the spread could minimize mortality impressively than using the vaccines selecting elderly people. Tom Britton et al. in [7] developed an age-structured mathematical model according to social activity level. They estimated that if the basic reproduction number reaches 2.5, then the disease-induced herd immunity level could reach 43%, which is lower than the classical herd immunity level. Mukandavire et al. used a mathematical model fitted to COVID-19 daily new cases in South Africa to estimate the critical vaccination coverage that control the disease for different vaccine efficacy levels. They estimated that a vaccine with 70% efficacy can eradicate COVID-19 disease in south Africa [36]. In [17], Enahoro A. et al. used of a mathematical model to evaluate the effect of an imperfect COVID-19 vaccine on the eradication of COVID-19 disease in the United States. Their study suggests that the likelihood of COVID-19 eradication in the United States can be highly improved if the vaccination is accompanied with non pharmaceutical measures.

In this work, we develop a discrete age-structured model with a booster vaccination, focusing on an established path of COVID-19 transmission from children to adults and vice versa. In other words, we present a model, splitting the population into two different cohorts, to focus our investigation on a vaccination program targeting adults aged over 18 years. Since the developed COVID-19 vaccine may not always hold when carried out to the patient and doesn’t always protect against infection [21, 27], our model will include varying efficacies and immunogenecities of the vaccine against COVID-19. The model is a useful tool, which will be used to assess the vaccination rate, efficacy and the immunogenicity of vaccine needed to eradicate the COVID-19 according to the intended strategy followed by the Moroccan Government.

## 2. Method

### 2.1. Data collection

The data of reported symptomatic infectious and death cases is collected each day at 11 pm from the official Coronavirus Portal of Morocco [48]. Data information, as shown in Fig. 2.2 and Fig. 2.1 covers the cumulative number of reported cases and death cases from March 2*nd* to December 26*th*, 2020.

**Figure 2.1:**
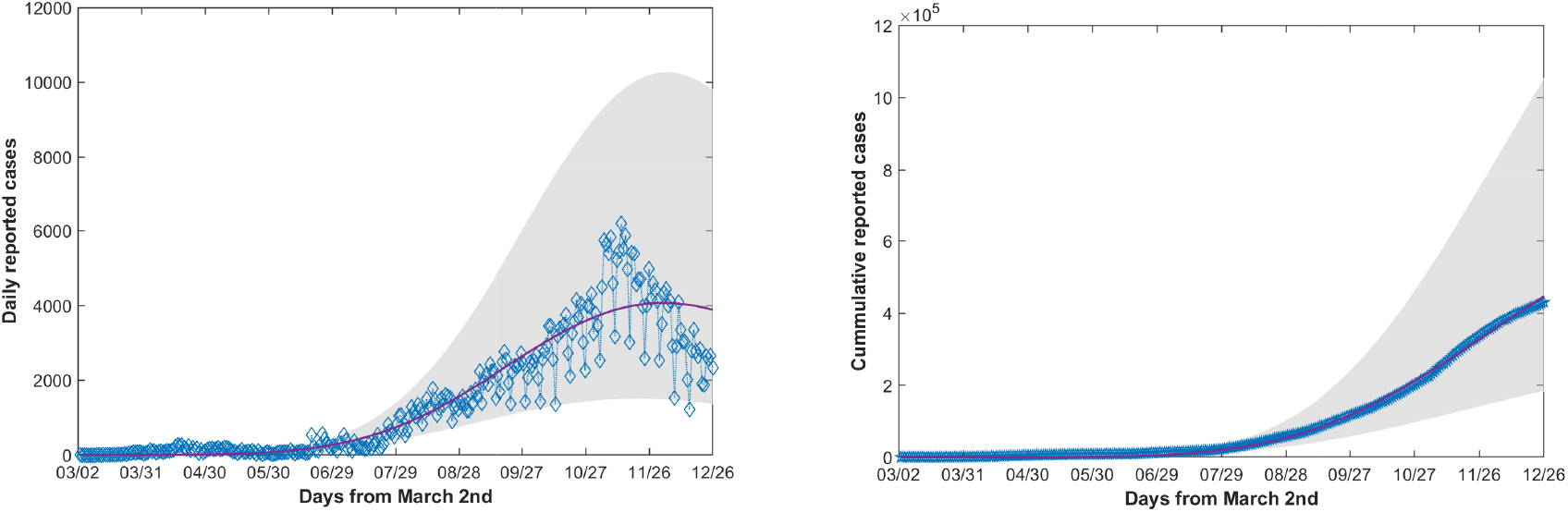
COVID-19 epidemic data and fitting results in Morocco using model (2.1) in the absence of vaccination between March 2*nd*, 2020 and December 26*th*, 2020.

**Figure 2.2:**
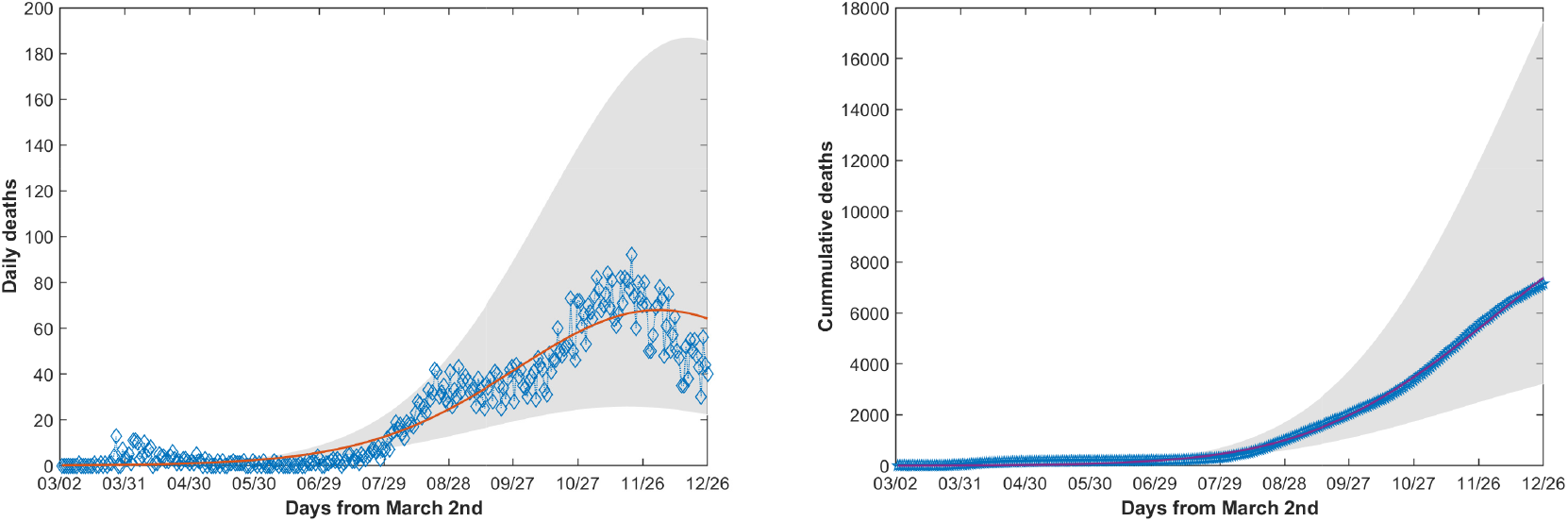
COVID-19 mortality data and fitting results in Morocco using model (2.1) in the absence of vaccination between March 2*nd*, 2020 and December 26*th*, 2020.

### 2.2. Model formulation

The population considered in this section is stratified into two age categories and eight disease status. Individuals are classified as unvaccinated susceptible children (*T*_*u*_), unvaccinated susceptible adults (*S*_*u*_), vaccinated susceptible adults (*S*_*v*_), asymptomatic infectious adults (*A*), asymptomatic infectious children (*B*), unreported symptomatic infectious (*I*_*u*_), hospitalized symptomatic infectious (*H*), recovered individuals (*R*) and COVID-deceased individuals (*D*). We assume that infected children do not show symptoms and can still transmit the disease. COVID-19 disease dynamics can be described as follows: Susceptible children are born at rate *π*_*T*_ and move to the susceptible adult class at rate *α*. Unvaccinated susceptible individuals that are vaccinated at rate *p* and acquire immunity from the vaccine at rate *ϵ* move to the vaccinated class (*S*_*v*_). It is assumed that vaccination does not necessarily induce lifelong immunity. Thus, vaccinated susceptible individuals are also assumed to acquire COVID-19 infection with a protective efficacy *ψ*, following effective contact with asymptomatic or unreported infected individuals. Furthermore, it is assumed that vaccinated class (*S*_*v*_) receives a booster dose as a second administration of the vaccine, at a rate *ϕ*, to maintain successful immunization. Unvaccinated susceptible adults (*S*_*u*_) (resp. vaccinated susceptible adults (*S*_*v*_), susceptible children (*T*)) are infected through contact with infectious adults (*A* + *I*_*u*_) at a transmission rate *β*_*aa*_ (resp. (1 − *ψ*) *β*_*aa*_, *β*_*ca*_) or through contact with infectious children (*B*) at a transmission rate *β*_*ac*_ (resp. (1 − *ψ*) *β*_*ac*_, *β*_*cc*_). Once infected, children and adults move, respectively, to the asymptomatic infectious class (*B*) and the asymptomatic infectious class (*A*). After an average period 1*/δ* days the asymptomatic infectious individuals (*A*) become symptomatic and proceed either to the unreported symptomatic infectious (*I*_*u*_), at rate *δ*_1_, or to the hospitalized individual (*H*) at rate *δ*_2_ with *δ* = *δ*_1_ +*δ*_2_. Once becoming symptomatic, individuals of class *I*_*u*_ and *H* either remain asymptomatic for 1*/µ* days on average before they are recovered or remain asymptomatic for 1*/d* days on average before they are dead due to infection. Asymptomatic children can either be recovered without being hospitalized at rate *δ* or detected and hospitalized at rate *σ*. All individuals are naturally dead at rate *d*_*N*_. The model will be given by the following equations

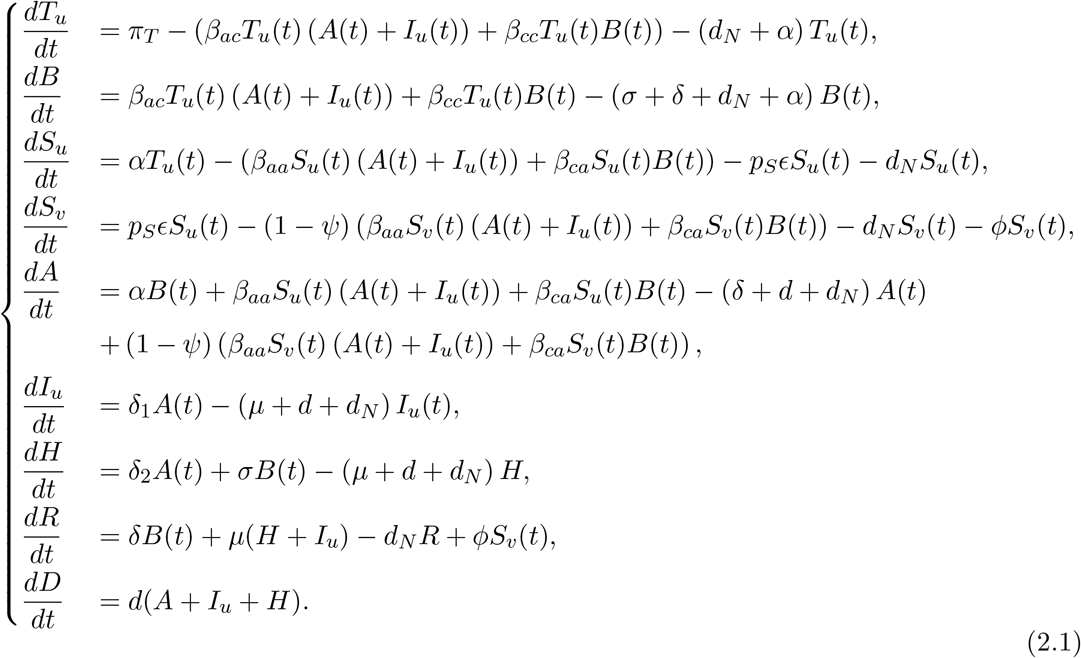

### 2.3. Parameter and Initial Data Estimation

To estimate the model parameters we will use the data between March 2*nd* and March 20*th*,2020 during which there were no control measure. We will assume that *S*_*v*_ = 0, *p*_*S*_ = 0 and *ϵ* = 0. Note that the first infected child under 18 years was reported 22 days since the beginning of the epidemic. Furthermore, the maximum asymptomatic duration is assumed to be around 6 days. Consequently, it is meaningful to assume that there were no asymptomatic infected children (i.e. *B*(0) = 0) under 18 years old at time *t* = 0.

The cumulative reported infectious population is given, for *t* ≥ 0, by 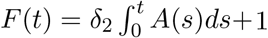. It is obvious that cumulative reported infectious population increases slowly and then accelerates rapidly with time. Hence, we will use exponential regression with 95% of confidence level to find an exponential function that best fits the data, from March 2*nd* to June 10*th*. Using SPSS software (Statistical Package for the Social Sciences) we found that the exponential model, given by *be*^*at*^ where *a* = 0.263 with confidence interval *CI* (0.229 − 0.297) and *b* = 0.507 with *CI* (0.3444 − 0.7475) fits well the data with a correlation coefficient given by *R* = 0.97. It follows from 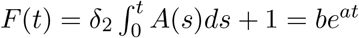 that

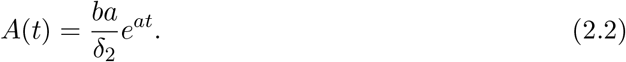

Since the initial susceptible population is not dramatically affected in the early phase of the epidemic, we will assume that *S*_*u*_(*t*) *≈ S*_*u*_(0). Let *S*_0_ := *S*_*u*_(0), *A*_0_ := *A*(0) and *I*_0_ := *I*_*u*_(0). From the fifth equation of system (2.1) and using (2.2) we obtain

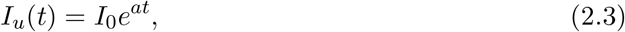

where

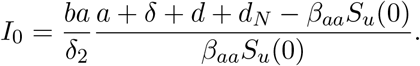

Now, using formulas (2.3) and the sixth equation of system (2.1), we obtain after simplification

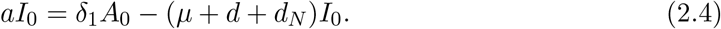

It follows that

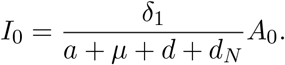

Furthermore, *S*_*v*_(0) = 0, *T*_*u*_(0) = 12437000 and *S*_*u*_(0) = 23428191. We calculate the leaving rate of children under 18 years old to adults as 1*/α* = 18 *×* 365 days, thus, *α* = 1, 52 *×* 10^−4^. Since the Moroccan authorities decided to give the second dose of the vaccine after 21 days following the first dose then we assume that *ϕ* = 1*/*21. As mentioned in Section 1, we assume that *β*_*ac*_ = *β*_*aa*_ and *β*_*ca*_ = *β*_*cc*_. To estimate the transmission rates, *β*_*aa*_, *β*_*cc*_ and *d*, we use the nonlinear least squares solver “lsqcurvefit” in MATLAB R2019b software. The values of the estimated parameters are summarized in Table 1.

**Table 1:** Parameter definitions and values of model (2.1).

| Symbol | Definition | Parameter value | Confidence interval | Reference |
| --- | --- | --- | --- | --- |
| $S_u(0)$ | Initial susceptible population | 23428191 | | [20] |
| $T_u(0)$ | Initial susceptible children | 12437000 | | [20] |
| $S_v(0)$ | Initial vaccinated population | 0 | | See text |
| $A(0)$ | Initial asymptomatic population | 0.9 | 0.48 – 1.357 | [19] |
| $I_u(0) (\times 10^{-2})$ | Initial unreported symptomatic population | 6 | 2.9 – 8.3 | [19] |
| $H(0)$ | Initial reported symptomatic population | 1 | | See text |
| $R(0)$ | Initial recovered population | 0 | | See text |
| $D(0)$ | Initial COVID-deceased population | 0 | | See text |
| $\beta_{aa} (\times 10^{-9})$ | Infection rate from infectious adults to individuals | 8 | 6.82 – 10.77 | Estimated |
| $\beta_{cc} (\times 10^{-9})$ | Infection rate from infectious children to individuals | 1.14 | 0.97 – 1.5 | Estimated |
| $\alpha (\times 10^{-4})$ | Leaving rate of children under 18 years old | 1, 52 | | Calculated |
| $1/\delta$ | Asymptomatic duration | 6 days | | [48] |
| $\delta_1$ | Asymptomatic unreported rate | 0.017 per day | | Assumed |
| $\delta_2$ | Symptomatic reported rate | 0.15 per day | | Assumed |
| $1/\mu$ | Symptomatic duration | 14 days | | [58] |
| $p_S$ | Proportion of primary vaccination under booster administration | 0 – 1 | | Varied |
| $d$ | Death rate from infection | 0.16 | | Estimated |
| $\epsilon$ | Immunogenicity | 0 – 1 | | Varied |
| $\psi$ | Efficacy of the vaccine | 0 – 1 | | Varied |
| $\phi$ | Rate of second dose of vaccine | 1/21 | | Calculated |
| $\pi_T$ | Birth rate | 1572 | | [20] |
| $d_N (\times 10^{-5})$ | Natural death rate | 3.34 | | [20] |

## 3. Results

Since the three last components *H,R* and *D* do not appear in the six first equations of model (2.1) then we will focus our local stability study on the following system.

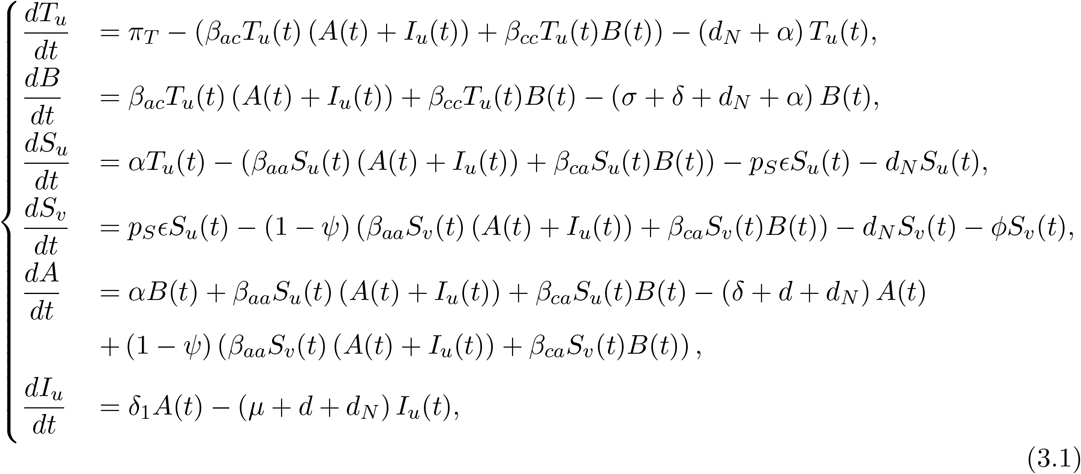

It is obvious that the model (3.1) has a unique disease free equilibrium (DFE) which is given by 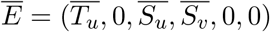, where

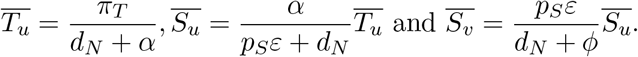

### 3.1. Control reproduction number

The control reproduction number, *R*_*c*_, is an important value, used to determine whether a control policy, such as lockdown, lifting, behavioral practices, vaccination, etc, will be efficient to decrease the number of secondary infections to be less than one. In the absence of any control, this number is considered as the basic reproduction number, *R*_0_, which is the average number of secondary infections produced when one infectious individual is introduced into a host susceptible population. This quantity determines whether a given disease may spread, or die out in a population. Applying the next generation matrix method formulated in [51], *R*_*c*_ can be given by

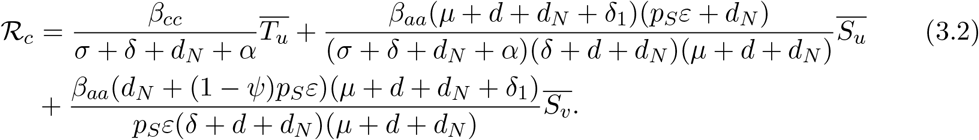

The calculation of *R*_*c*_ can be found in Appendix A.

### 3.2. Dynamics of the disease-free equilibrium

The local behavior of the DFE of system (3.1) is given by the following theorem.

#### Theorem 1.

*The disease-free equilibrium is locally asymptotically stable for ℛ*_*c*_ *<* 1 *and unstable for ℛ*_*c*_ *>* 1.

The proof of Theorem 1 can be found in Appendix B. The next lemma will be useful to prove the global stability of the DFE.

#### Lemma 2.

*The region*

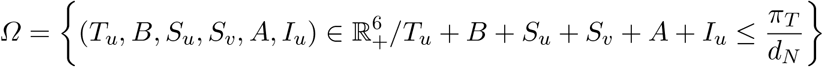

*is positively invariant and attracting with respect to system ((3.1))*.

The proof of Lemma 2 can be found in Appendix C.

Put

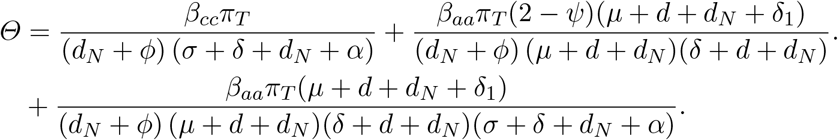

Now we are in the position to state and prove the global asymptotic stability of the DFE.

#### Theorem 3.

*Assume that Θ <* 1 *and ℛ*_*c*_ ≤ 1, *then the disease free equilibrium E of system ((3.1)) is globally asymptotically stable in the region Ω, i.e*,

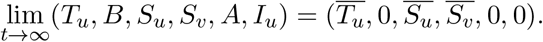

The proof of Theorem 3 can be found in Appendix D.

#### Remark 4.

Using parameter values in Table 3 and varying the vaccine efficacy values, *ψ*, in the range [0, 1], one can show that the parameter values of *Θ* vary in the range [−0.9799, −0.9788]. It follows that, in order to assure the global asymptotic stability, it suffices to assume that *ℛ*_*c*_ ≤ 1.

### 3.3. Herd immunity

Herd immunity is defined as a level of population immunity at which the amount of virus able to spread in the entire population becomes very low so that the spread of the disease will be over. However, if the immunity level does not reach the herd immunity level, then a second wave of disease spread may appear. Thus, it is an interesting to look when herd immunity can be reached. Although natural herd immunity is considered as a way to reach herd immunity through natural recovery from COVID-19 infection, vaccination is considered as the best way to protect susceptible individuals and their community. Indeed, once plenty individuals are vaccinated and get immune from the disease, viruses cannot pass between individuals in the population and the whole community is less probably to contract the disease.

Note that the quantity *ϕ*, given in (A.1), can be written as

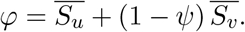

Thus, *ℛ*_*c*_ will be given by

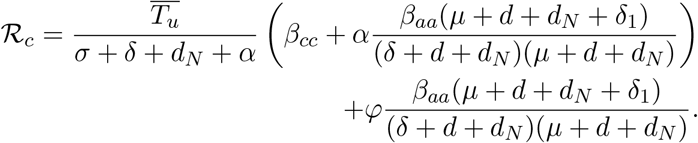

So *ℛ*_*c*_ = 1 is equivalent to

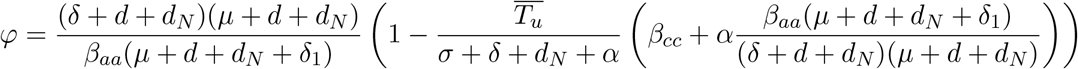

On the other hand, we have 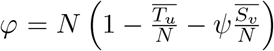, where *N* is the total population size. Let 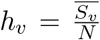 and 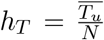, respectively, be the the proportions of susceptible individuals that have been vaccinated and susceptible children at the disease-free equilibrium. Set

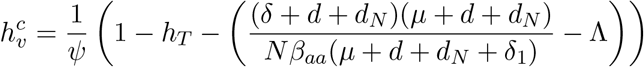

where

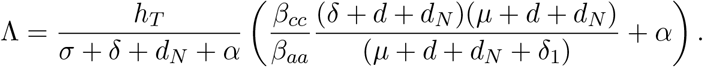

Note that, following Theorem 3 and Remark 4, the disease free equilibrium 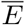 of system ((3.1)) is globally asymptotically stable if *ℛ*_*c*_ *<* 1. Furthermore, *ℛ*_*c*_ *<* 1 if and only if 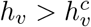 and, thus, for the herd immunity to be achieved, a proportion 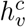, which is known as the herd immunity threshold, of the entire population should be vaccinated. Furthermore, if the vaccination is not successful at all (*ψ* = 0) then no matter the proportion of vaccinated adults, the eradication of the COVID-19 disease is implausible.

## 4. Simulations and discussion

The vaccination is among the most effective methods of preventing disease, disability, and death. A vaccine is defined as a suspension of live attenuated or inactivated pathogens, or fractions thereof, which is administered to induce immunity and prevent infectious disease or its sequelae [46]. Throughout 2020, a global effort have been undertaken to produce safe and effective vaccines against the newly emerged coronavirus, SARS-CoV-2. Actually, several of them received approval to initiate the national and multi-country mass vaccination campaigns. Morocco also will launch an emergency vaccination program for the adult population using vaccines produced by pharmaceutical firm Sinopharm and Astrazeneca. Over a period of 3 months, susceptible adult people who volunteer for the vaccination will receive two injections of vaccine spaced by 21 days apart [49]. Generally, the first dose induces a primary immune response by exposing the body for the first time to antigens. This immune response is often slow, limited and do not lead to protective immunity [45]. Therefore, a subsequent dose, also called a booster shot, is required to induce a secondary immune response in order to maintain an immunological memory which provides a long-time immunity. Indeed, the long-lived antibodies and memory cells have the ability to respond faster and more efficiently to the re-exposure to the same antigens [1]. Unfortunately, it is still too early to assess the duration of immunity provided by all anti-COVID-19 vaccines [3, 9]. Moreover, our understanding to the magnitude and duration of the immune response to natural infection with SARS-CoV-2 remains limited to date [12]. But it is well known that, for the common coronaviruses, the protective immunity is short-lasting and the reinfection often occurs 12 months after infection [8, 16]. For the SARS-CoV-2, several studies have shown that the immunity wanes over time and can be maintained only for few months after infection [54, 25, 44, 14]. Furthermore, the report of many cases of secondary infections with the COVID-19 in the same year also provides evidence that the acquired immunity is short-lived [16]. In case the protective immunity, conferred by the infection, is transient like for the coronavirus diseases, it will be necessary to plan periodic vaccination campaigns in coming years to prevent future outbreaks [40, 12].

In this study, we used a discrete age-structured mathematical model to assess the SARS-CoV-2 vaccination program adopted by Morocco incorporating the efficacy and immunogenicity of the used vaccine and the vaccination rate of adult population. The data used for simulations include implicitly the effect of the NPI (i.e. wearing masks; social distancing; cleaning and disinfecting measures…) applied between March 2nd, 2020 and December 26th, 2020 on COVID-19 disease spread. Based on our results, the COVID-19 is likely to persist past 2021 in the absence of a vaccine. Indeed, the number of daily reported cases and deaths tend to decline slowly over time, reaching, respectively, a number of 800 and 10 cases per day by the end of 2021 (See Fig. 4.1 and Fig. 4.2).

**Figure 4.1:**
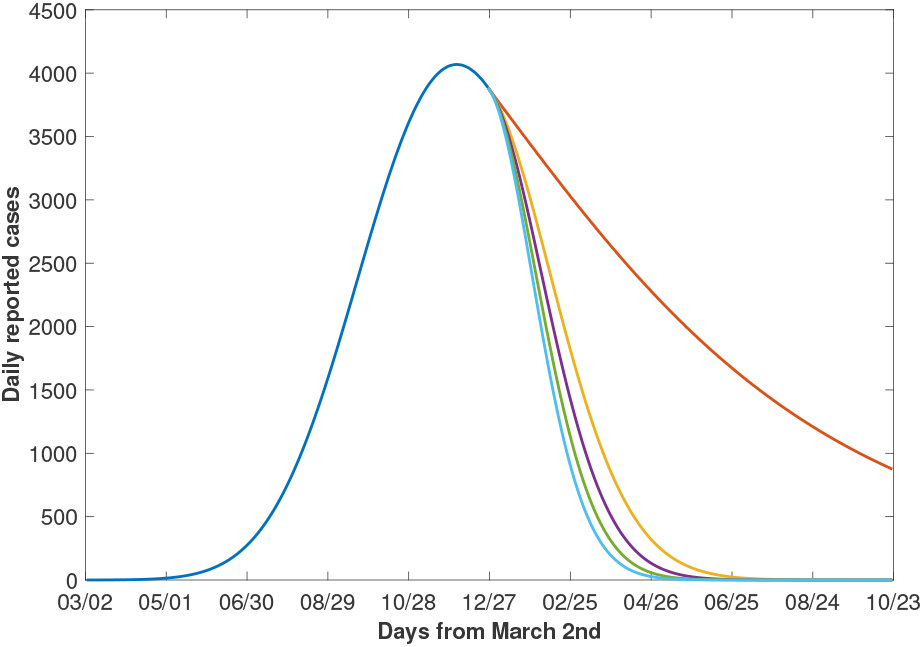
Daily reported cases over time. Daily reported cases are shown when vaccination is given with levels, from top to bottom, 0, 0.5, 0.6, 0.7 and 0.8% per day. Vaccine efficacy (*ψ*) is assumed to be 90 and the immunogenicity (*ϵ*) is assumed to be 50%.

**Figure 4.2:**
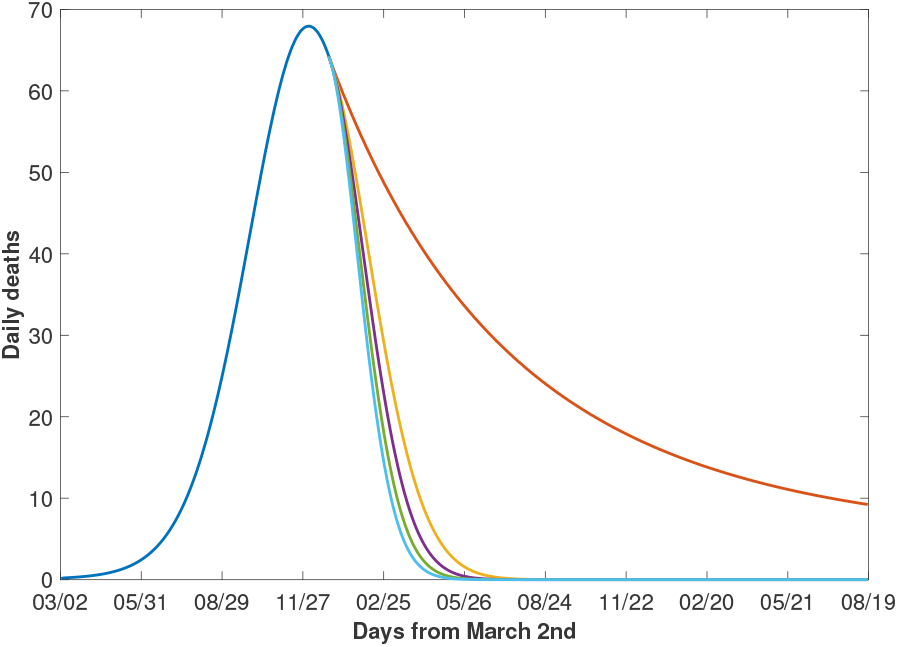
Daily death cases over time. Daily mortality cases are shown when vaccination is given with levels, from top to bottom, 0, 0.5, 0.6, 0.7 and 0.8% per day. Vaccine efficacy (*ψ*) is assumed to be 90 and the immunogenicity (*E*) is assumed to be 50%.

However, although it takes many months, vaccinating adults could help to eradicate the disease more quickly. The Moroccan people will have to wait until the second half of 2021 before definitively abandoning the NPI. Several studies have shown that when the NPI are maintained during the vaccination program, the COVID-19 elimination is greatly improved in the country such as the US and South Africa [24, 4, 36].

They also found that, in the absence of NPI, the vaccines with at least 70% efficacy are able to contain COVID-19 outbreak but higher vaccination coverage is needed. In the case of Morocco, the efficacy and immunogenicity of the vaccine needed to eradicate the COVID-19 depend on the proportion of adults vaccinated daily. Indeed, if the Moroccan government plans to immunize the adult population at a rate of 0.6% per day, then the used vaccine must have an efficacy rate of at least 67% and generate a strong immunogenicity (*E* = 0.9) in vaccinated individuals to achieve the disease eradication (Fig. 4.3). The use of UK’s Oxford-Astrazeneca or China’s Sinopharm vaccine that have, respectively, a efficacy rate of 70% or 79% in vaccinating the adult population may be powerful tool in the COVID-19 eradication while still maintaining the NPI [28]. However, although difficult to enact, if the Moroccan health care is able to increase the vaccination rate to 1% per day, then vaccine has to have at least 40% of efficacy to eradicate the disease (Fig. 4.3). So the success of the vaccination program depends not only on the efficacy rate of vaccine and the degree of the generated immune response but depends also on the logistical resources deployed to ensure vaccination of large numbers of individuals within a short space of time. Paltiel and its collaborators [41] also showed that the advantages of a vaccine can decline significantly in the event of manufacturing or deployment delays, significant vaccine hesitancy, or greater epidemic severity. Many pharmaceutical companies have taken up the challenge of supplying the world with the first anti-COVID-19 vaccines at the end of 2020. However, they are now facing a new challenge to produce a sufficient quantity of COVID-19 vaccines of high quality to cover the demand of all emerging and non-emerging countries affected by this pandemic. According to scientific news published by Cohen and Kupferschmidt in [12], it will be necessary to wait for the year 2022 to assure enough vaccines available to vaccinate people wishing to be vaccinated. Fortunately, reaching the so-called herd immunity threshold could have an indirect effect on the protection of the unvaccinated susceptible people against the infection. The notion of herd immunity threshold is defined as the proportion of immunized individuals, through natural infection or through vaccination, in a population who cannot get infected anymore [40]. Generally, although herd immunity threshold varies with each disease, a large proportion of individuals must be immunized to achieve the herd immunity. For the COVID-19, the herd immunity threshold is still unknown and will probably vary depending on the community and the vaccination strategy adopted by the country in question among others [62]. However, several studies using mathematical modeling were able to estimate this threshold [24, 4, 36]. For example, by assuming a vaccine with 80% of efficacy, Iboi and collaborators [24] found that at least 82% of susceptible population in US must be immunized against the SARS-CoV-2 to achieve the herd immunity threshold. Moreover, they also found that this percentage could drop to 72% (resp. 46%) with a regular wearing of masks by the half of the population (resp. the entire population). In our case, the herd immunity is achieved when about half of Moroccan adult population is immunized against the COVID-19 with a vaccine efficacy rate of 79% like the one by Sinopharm vaccine (See Figure 4.4). However, with an efficacy rate of 70% such as the case of Oxford-Astrazeneca vaccine, less than 60% of adult population must be immunized against the disease to achieve the herd immunity.

**Figure 4.3:**
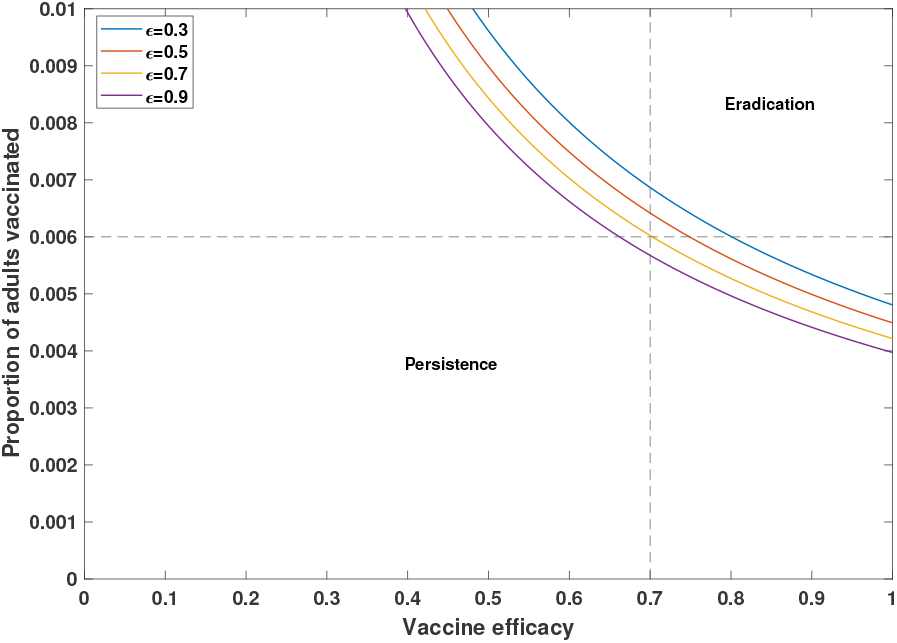
Vaccination thresholds for eradication depending on vaccine efficacy for the four levels of immunogenicity, from top to bottom, 30%, 50%, 70% and 90% at the time of vaccination. The vertical line corresponds to *ψ* = 0.7 while the horizontal line corresponds to *p*_*S*_ = 0.006.

**Figure 4.4:**
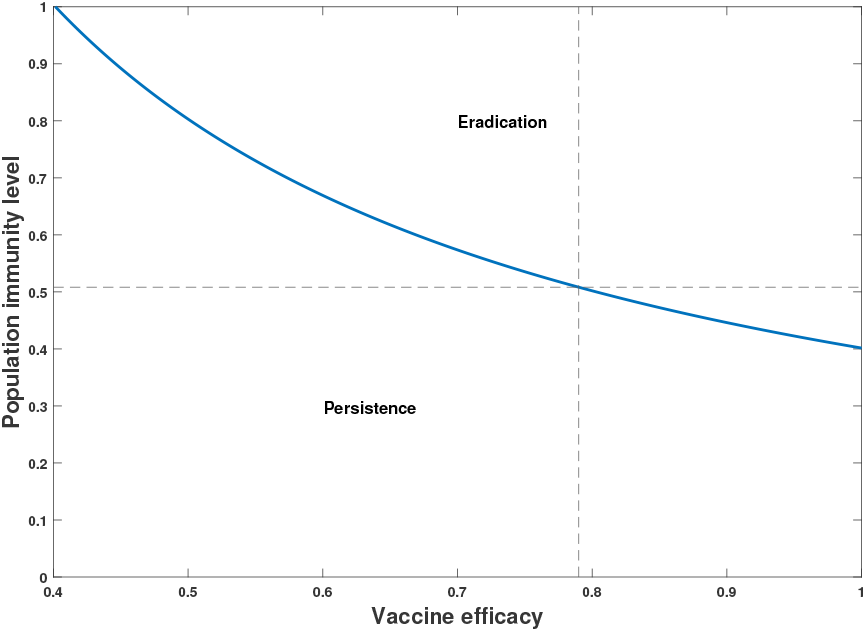
Population immunity threshold for eradication depending on vaccine efficacy at the time of vaccination. The vertical line corresponds to *ψ* = 0.79 while the horizontal line corresponds to *h*_*v*_ = 0.508.

These proportions should probably be higher than estimated since our model also takes into account the combined effects of the NPIs and vaccine.

Finally, note that COVID-19 vaccines obviously provide direct protection to adults but can also produce indirect protection and block the course of infection to children. The Fig. 4.5 shows that, if vaccine efficacy is about 80% and the immunogenicity is about 50%, then protected susceptible children increases as vaccine uptake increases in the total adult population. In particular, 0.6% of vaccinated adults could save about 22% of children from infection.

**Figure 4.5:**
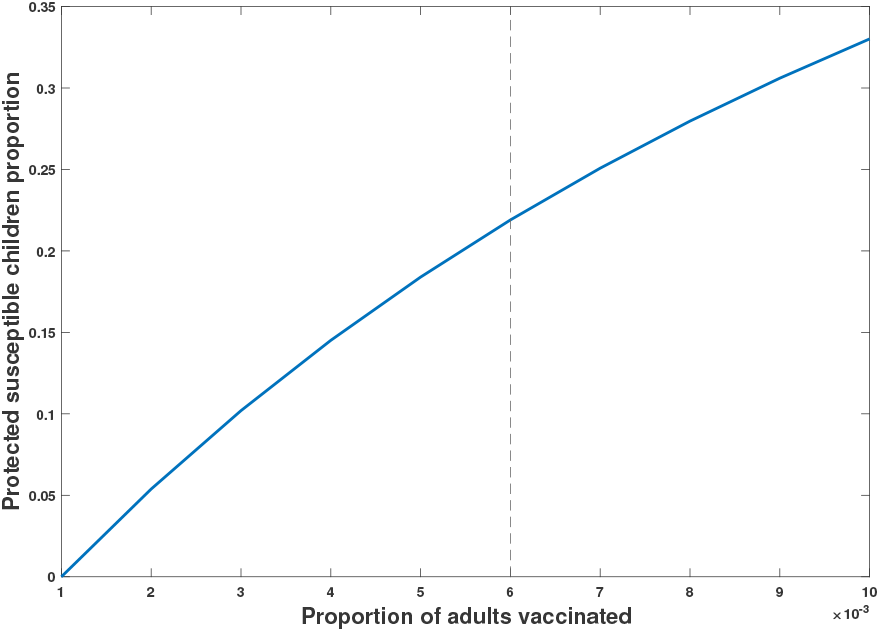
Proportions of protected susceptible children with different levels of vaccination. Proportions of protected susceptible children increases as vaccine uptake increases in the total adult population. Vaccine efficacy (*ψ*) is assumed to be 80 and the immunogenicity (*ϵ*) is assumed to be 50%. The vertical line corresponds to *p*_*S*_ = 0.006. (Protected are 22% of susceptible children)

## 5. Conclusion

While waiting for the first doses of vaccines since the end of December 2020, Morocco is preparing for the vaccination campaign of the adult population to eradicate the COVID-19 epidemic. Our findings show that, with a vaccination rate of 0.6% per day, the vaccine must both have an efficacy rate of at least 67% and must provide a high degree of immunity to people vaccinated. In addition, it is also necessary to continue to take protective measures during the vaccination period to achieve the herd immunity threshold more quickly. Our results found that the herd immunity can be obtained by immunizing less than 60% of the adult population against the COVID-19 with a vaccine with 70% or more of efficacy. Finally, this vaccination program can also protect 22% of susceptible children from infection. However, in order not to devalue the efficacy of the vaccine used and before starting vaccination campaign, the Moroccan government must ensure the availability of all doses of vaccine needed to cover the entire target population, encourage the concerned people to be vaccinated, and carry out all vaccinations in the shortest time in accordance with the human, logistical and financial resources of country.

## Data Availability

Yes

http://www.covidmaroc.ma

## Acknowledgments

The authors thank the anonymous referees, whose careful reading, insights, valuable comments, and suggestions significantly enabled us to improve the quality of the paper.

## Appendix A

Following the notation in [51], the associated next generation matrices, *F* and *V*, for the new infection terms and the transition terms, are given, respectively, by

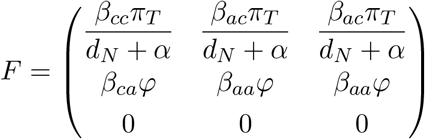

and

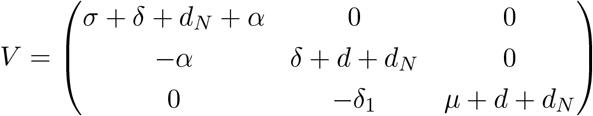

Where

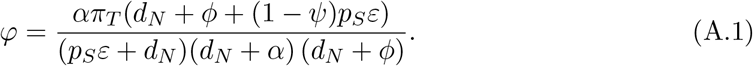

We have det(*V*) = (*σ* + *δ* + *d*_*N*_ + *α*)(*δ* + *d* + *d*_*N*_)(*µ* + *d* + *d*_*N*_). Then, the inverse of *V* is given by

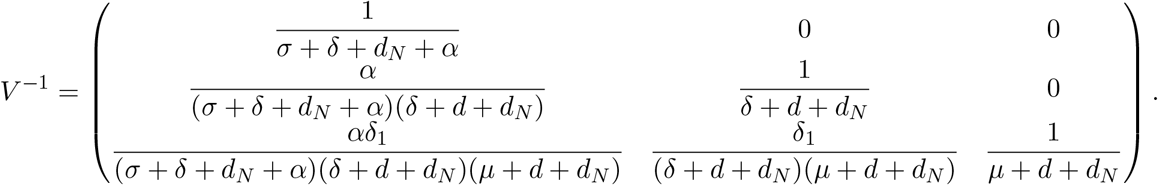

Thus, the next generation matrix for system ((3.1)) will be given by

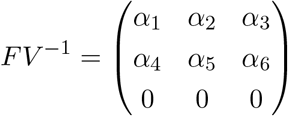

Where

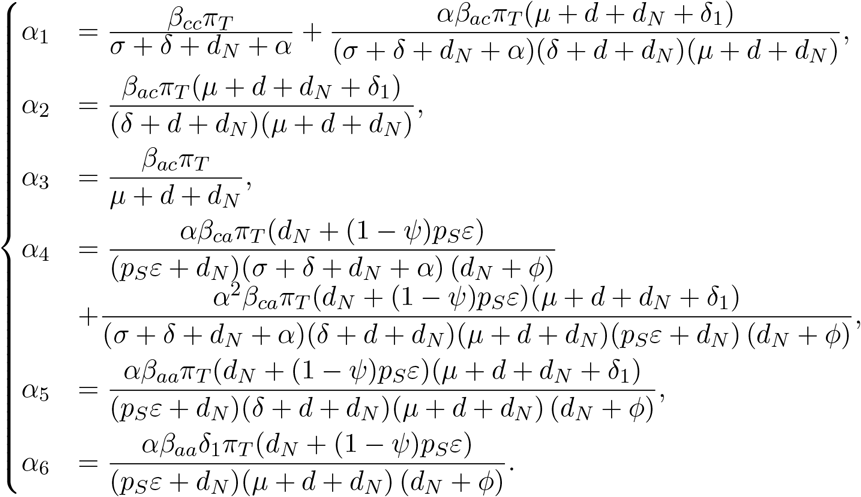

Therefore the associated characteristic equation is given by

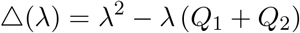

where

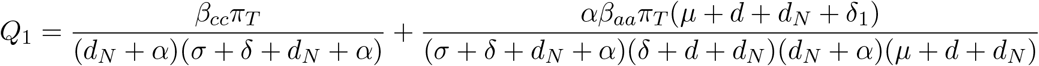

and

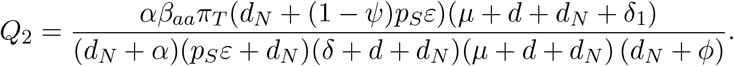

Therefore, by definition of *ℛ*_*c*_, we obtain

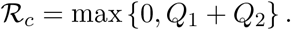

Consequently, the control reproduction number is given by formula (3.2).

## Appendix B

The local stability of 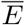 is governed by the eigenvalues of the Jacobian matrix, of system ((3.1)),

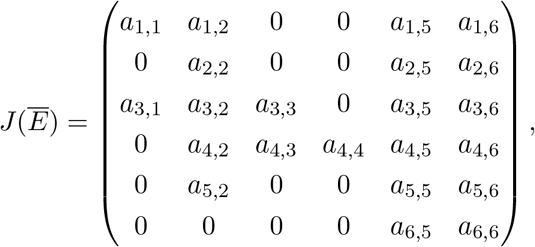

Where

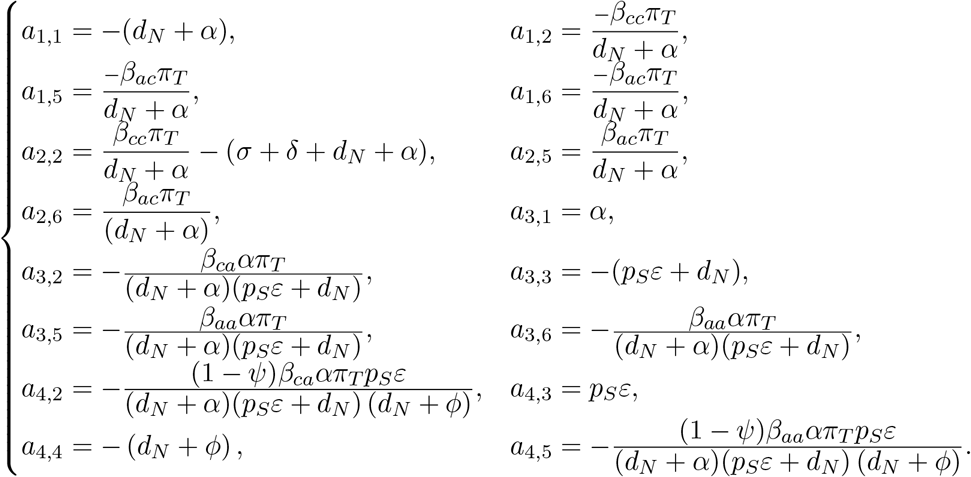

and

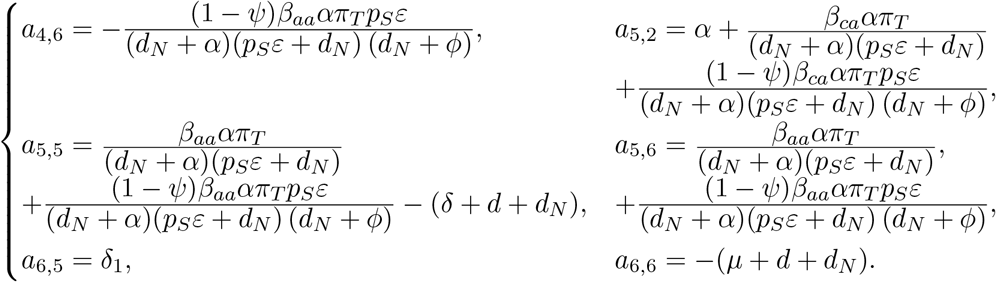

Therefore the associated characteristic equation is

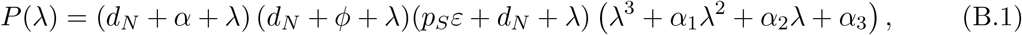

where

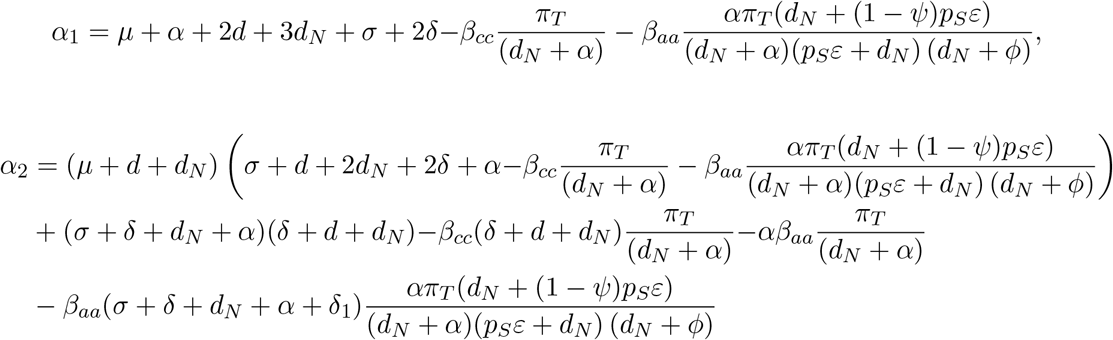

and

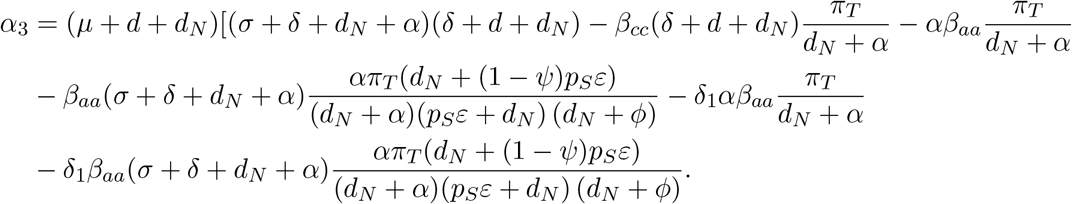

Then −(*d*_*N*_ + *ϕ*), − (*p*_*S*_*ε* + *d*_*N*_), − (*d*_*N*_ + *α*) are roots of the characteristic equation (B.1) and, consequently, a necessary and sufficient condition for the other roots ta have negative real parts is given by the following lineard-chipart [32] criteria

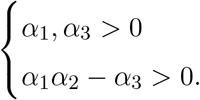

On can see that

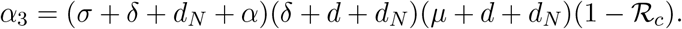

Moreover, if *ℛ*_*c*_ *<* 1, then

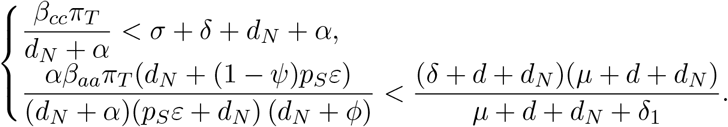

It follows that

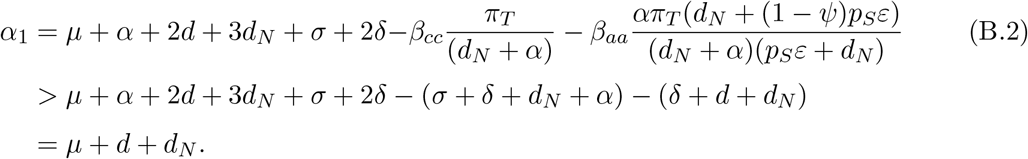

Thus, if *ℛ*_*c*_ *<* 1 then *α*_1_ and *α*_3_ are positive. Now let’s prove that *α*_1_*α*_2_ − *α*_3_ *>* 0. After few simplification we can get

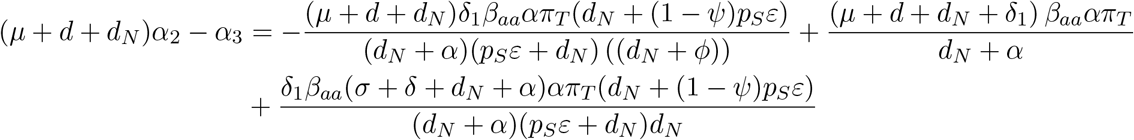

and, consequently,

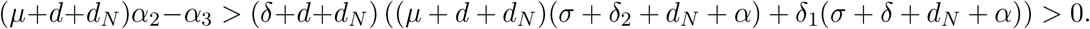

Furthermore, from ((B.2)), we have *α*_1_ *> µ* + *d* + *d*_*N*_. Then, *α*_1_*α*_2_ *>* (*µ* + *d* + *d*_*N*_)*α*_2_ *> α*_3_. Finally, we have *α*_1_*α*_2_ − *α*_3_ *>* 0. Hence, *E* is locally asymptotically stable for *ℛ*_*c*_ *<* 1.

Assume now that *ℛ*_*c*_ *>* 1. Since

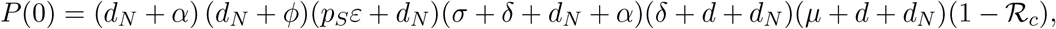

then *P* (0) *<* 0. Moreover, when λ ∈ ℝ then 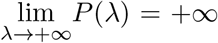, Then, there exists *λ*_0_ *>* 0 such that *P* (*λ*_0_) = 0 and the disease-free equilibrium becomes unstable. This completes the proof.

## Appendix C

For the invariance property it suffices to show that the vector field, on the boundary, does not point to the exterior. On one hand, on the boundary of *Ω*, we have

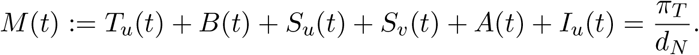

Then

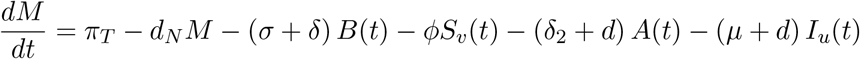

and, consequently,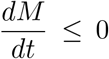. Therefore, solutions starting from *Ω*, will remain there for *t* ≥ 0. On the other hand, we have 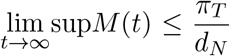. Hence, *Ω* is attracting, that is, all solutions of ((3.1)) eventually enters *Ω*. This completes the proof.

## Appendix D

To prove the above result, we use the following Lyapunov function

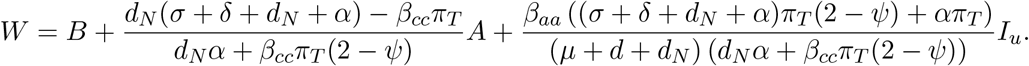

The orbital derivative of *W* is given by

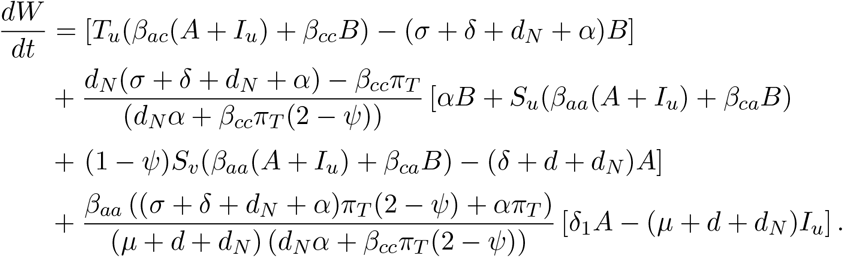

Since 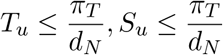 and 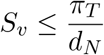, we obtain

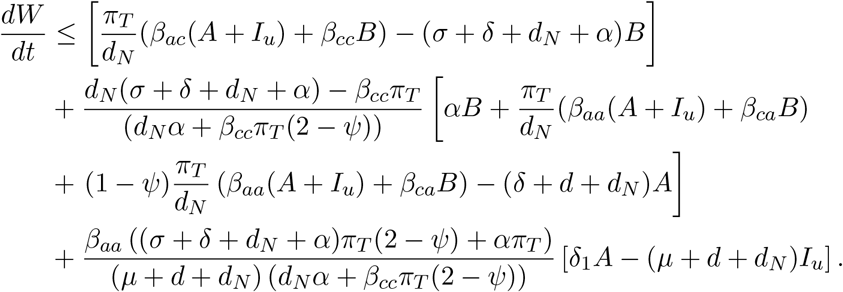

Thus,

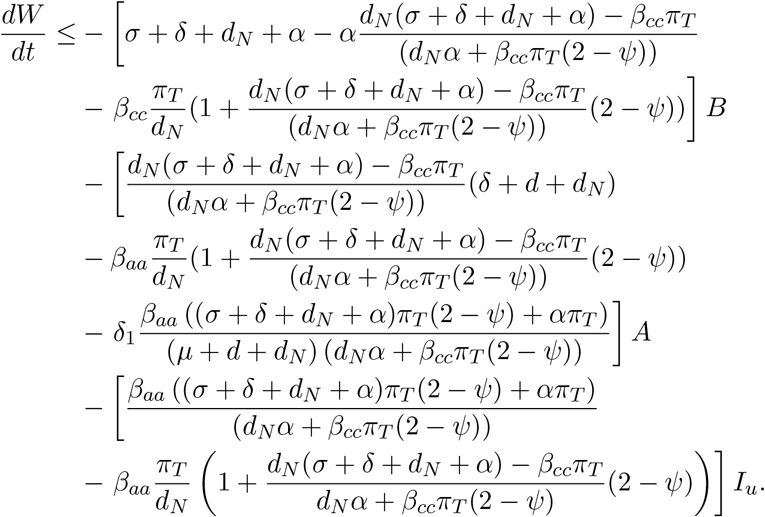

Notice that

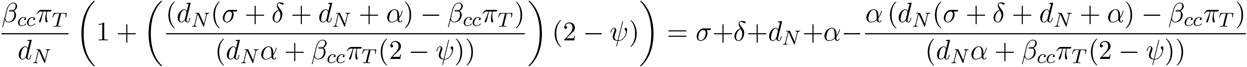

and

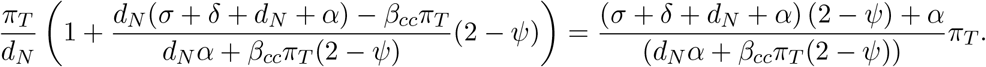

It follows that

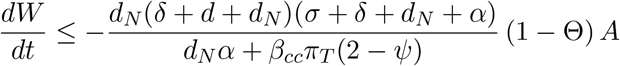

where

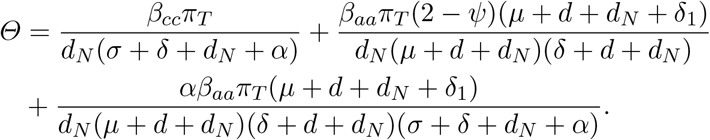

Consequently 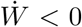 whenever Θ *<* 1. Furthermore, 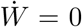 implies that (1 − Θ) *A* ≤ 0. It follows that *A* = 0 and, thus, the fifth equation of system ((3.1)) becomes

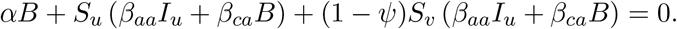

Then *B* = *I*_*u*_ = 0 and, consequently,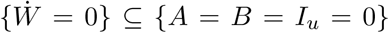. It follows that the maximum invariant set contained in 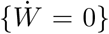. is the set *{A* = *B* = *I*_*u*_ = 0*}*. Any solution start from the set *{A* = *B* = *I*_*u*_ = 0*}* must satisfies

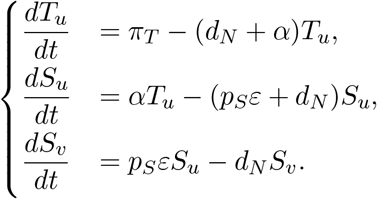

It follows that

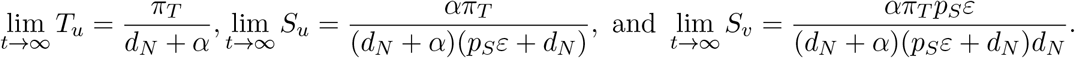

Therefore, applying the LaSalle–Lyapunov Invariance Principal, it follows that *E* is globally stable.

